# Outcomes of prognostication in people living with advanced cancer: a qualitative study to inform a Core Outcome Set

**DOI:** 10.1101/2024.02.08.24302518

**Authors:** Caitlin Spooner, Bella Vivat, Nicola White, Patrick Stone

## Abstract

**Background:** Studies of prognostication in advanced cancer use a wide range of outcomes and outcome measures, making it difficult to compare these studies and their findings. Core Outcome Sets facilitate comparability and standardisation between studies and would benefit future prognostic research. This qualitative study is the second step in developing such a Core Outcome Set, with the aim to explore the perceptions and experiences of patients with advanced cancer, informal caregivers, and clinicians regarding the potential outcomes of prognostication.

**Methods:** We conducted semi-structured interviews with patients living with advanced cancer (*n*=8), informal caregivers (*n*=10), and clinicians (*n*=10) recruited from palliative care services across three sites in London, United Kingdom. Interviews were conducted in-person, via telephone, or video conferencing, and were audio-recorded. Data were analysed using a thematic framework analysis approach. Findings were compared with outcomes derived from a previously published systematic review.

**Results:** We identified 33 outcomes, 16 of which were not previously reported in the literature. We grouped outcomes into 10 domains, using a modified COMET taxonomy: 1) mortality/survival; 2) general physiological/clinical outcomes; 3) psychiatric outcomes; 4) spiritual/religious/existential functioning/wellbeing; 5) emotional functioning/wellbeing; 6) social functioning; 7) delivery of care; 8) perceived health status; 9) personal circumstances; 10) societal/carer burden. These findings highlighted discrepancies between the priorities of existing research and those of stakeholders.

**Conclusions:** This study offers valuable insights into outcomes significant to key stakeholders, underscoring the need for a patient-centred approach in research and clinical practice in prognostication in advanced cancer. These outcomes will play a key role in the development of a Core Outcome Set to assess the impact of prognostication in advanced cancer.

## Introduction

Patients with advanced cancer often seek their survival estimates, as this knowledge affects their life and healthcare choices [1, 2]. However, end-of-life prognostication is inherently complex, with ongoing debates regarding the most effective methods for making these predictions. An ideal method of prognostication would not only be accurate, but also positively influence patient decision-making, outcomes, and associated healthcare costs, a concept known as the ‘impact’ of prognostication [3].

Recent literature has increasingly focused on the accuracy of prognostication in advanced cancer care [4–8]. However, there is a notable gap in understanding the broader impact of these prognostic methods. This is partly due to the lack of agreed outcomes for assessing the impact of prognostication, and the heterogeneity in outcomes and measures used, which poses challenges to inter-study comparability [9]. The Core Outcome Measures for Effectiveness Trials (COMET) initiative seeks to develop Core Outcome Sets (COS) to improve consistency and comparability between research studies in any specific area [10]. This approach could prove helpful for prognostic studies.

We are conducting a wider study to develop a COS for prognostic studies in advanced cancer [11]. The initial phase of this overall study involved a systematic literature review examining the outcomes used to measure the impact of prognostication in previous studies, which found considerable diversity in outcomes and measures used [9]. Furthermore, the review confirmed that outcomes that have been used in previous studies might not be meaningful to those most affected by the research, since the perspectives of key stakeholders, such as patients, informal caregivers, and clinicians, remain largely unexplored [12–14]. There is an increasing emphasis on incorporating the perspectives of service users and providers in healthcare outcome evaluation [15]. Consequently, a key aspect of COS development involves identifying outcomes that are meaningful to stakeholders, ensuring that the outcomes included in any COS align with their priorities [16]. Qualitative research methodologies are increasingly used in the early stages of COS development to identify outcomes that are relevant and important to stakeholders [17].

However, to the best of our knowledge, there are no existing publications that detail the joint experiences and perspectives of key stakeholders on the impact of prognostication in advanced cancer. Therefore, we conducted a qualitative study within the broader context of our COS development project, to explore the experiences of prognostication from the perspective of patients with advanced cancer, informal caregivers, and clinicians. The objective of this study was to identify any additional potential outcomes of prognostication to those identified in the literature [9].

## MATERIALS AND METHODS

### Study design

We conducted a qualitative study using semi-structured interviews with an interpretive approach. This approach acknowledges the subjective nature of participants’ viewpoints and acknowledges that personal perceptions are shaped within the context of one’s environment [18]. This paper follows the Consolidated Criteria for Reporting Qualitative Research (COREQ) guidelines [19].

### Setting

Participants were recruited from palliative care services at two hospitals and one hospice in London, United Kingdom. Ethical approval was granted by the London-Camberwell St. Giles Research Ethics Committee and Health Research Authority on 6^th^ September 2022 (reference 22/LO/0469).

### Participants

We interviewed people from three stakeholder groups: patients with advanced cancer, informal caregivers, and clinicians.

To participate in the study, individuals had to be at least 18 years old at the time of giving consent and proficient in both spoken and written English. Patient participants were eligible if they had a confirmed diagnosis of advanced cancer of any type and were receiving palliative care at any of the participating sites. Informal caregivers qualified for inclusion if they were family members, friends, or other individuals who provided unpaid assistance or support to someone with whom they had a personal relationship. Clinicians eligible for participation included registered healthcare professionals (physicians, nurses, or allied health professionals, employed within the palliative care services at participating sites, who routinely estimates and provides prognostic predictions as part of their usual professional duties.

Exclusion criteria encompassed individuals whose involvement in the research was deemed inappropriate, as assessed by their attending clinician (e.g., due to a physical or mental condition), or those lacking capacity to give informed consent. Additionally, clinicians who worked in palliative care entirely outside the United Kingdom were excluded to eliminate variations in practice that could introduce confounding factors into the research.

### Sampling and recruitment

We employed a purposive sampling technique, aiming for diversity of participant characteristics. The recruitment process involved multiple strategies, including displaying posters at participating sites, direct identification, and outreach by the clinical team at each site, engagement by a research team member (CS) in multidisciplinary team meetings, and snowballing. Eligible participants were provided with an invitation letter and participant information sheet. CS subsequently contacted those who agreed to take part in the study. Written or verbal (audio-recorded) informed consent was sought from all participants on the day of the interview.

Recruitment continued until data saturation was attained, defined as the point at which no novel codes emerged from the interviews, and adequate data had been gathered to fulfil the research goals and objectives [20, 21].

### Data collection

Data collection took place between January and September 2023. One-to-one interviews were conducted either in-person (at offices based within our university), via telephone, or video conferencing. Interviews were conducted by CS, a female PhD student who has training in qualitative research methods, and no personal connections to any of the participants or the recruitment sites.

The interviews were semi-structured, following a topic guide (S1-S3 Files) with open-ended questions focused on prognostication experiences and an emphasis on the outcomes of prognostication which participants considered particularly important. Interviews were audio-recorded on an encrypted device, transcribed verbatim, pseudonymised, and managed using NVivo software (V.12) [22]. Transcripts were not returned to participants for corrections, to minimise burden. Data saturation was confirmed by the absence of new outcomes in the final interviews across all stakeholder groups.

### Data analysis

CS carried out initial interpretation and analyses, in consultation with the research team, and with regular guidance. Analysis was conducted alongside data collection to establish data saturation for each participant group.

Data were analysed using the Framework Method [23]. This approach provides a structured yet adaptable framework for conducting qualitative thematic analysis. It proves especially beneficial in qualitative research where the goal is to identify themes by conducting comparisons both within individual cases and across multiple cases [23]. This was the aim of our study, which set to explore experiences of prognostication from the perspectives of patients, informal caregivers, and clinicians.

The Framework Method involves a five-stage iterative approach to qualitative analysis: 1) data familiarisation, where researchers immerse themselves in the data to gain a deep understanding; 2) identifying a thematic coding framework, which involves creating a coding structure based on the data; 3) indexing and refinement of the coding framework, where data is systematically coded and the framework is adjusted as needed; 4) charting, involving the organisation of coded data into a structured format for analysis; 5) mapping and interpretation, where the structured data is analysed to identify patterns, relationships, and draw conclusion [24]. Initially transcripts were read multiple times to gain a comprehensive understanding of the data. This facilitated the generation of preliminary codes that captured the essence of participants’ experiences. Following data familiarisation, a coding framework was established. This framework had already been developed during the analysis of prior qualitative studies [9] and was guided by the outcome domains of the COMET taxonomy [10]. We then refined and broadened this framework by incorporating themes that extended beyond the established domains. This ensured a thorough and comprehensive capture of the data. The initial version of this framework underwent a review by the research team, which provided additional insights and refinements.

After refining our coding framework, we systematically applied the thematic framework or ‘index’ across all transcripts. The interview data were coded line-by-line for outcomes, defined as any perceived effect (beneficial or detrimental, intentional, or unintentional) of prognostication that participants identified for patients, informal caregivers, clinicians, or health services. A framework matrix was developed to facilitate the organisation of data samples from patients, informal caregivers, and clinicians. This matrix enabled the consolidation and grouping of codes (outcomes) that were preliminarily considered similar, aligning them within the appropriate outcome domains of the COMET taxonomy.

Participants did not provide feedback on the findings, to minimise burden. However, the preliminary grouping of outcomes and domains were presented to a Patient and Public Involvement (PPI) advisory group, comprising patients, informal caregivers, and public representatives, for validation. The research team then reviewed the provisional outcomes and their categorisation to ensure uniform interpretation. Disagreements were discussed until consensus on the final list of outcomes was achieved. Each outcome domain was summarised narratively, using illustrative extracts from the interviews.

Finally, we compared the identified outcomes against those found in a systematic review of outcomes previously reported in prognostic research with people with advanced cancer [9].

### Patient and public involvement

The wider COS project includes a PPI advisory group, composed of patients, informal caregivers, and public representatives. This group is involved in design, conduct, and dissemination of the whole project, including the analysis and interpretation of the results of this study.

## Results

### Participant characteristics

A total of 28 individuals (8 patients, 10 informal caregivers, and 10 clinicians) took part in interviews: 9 in person; 6 on the telephone; and 13 via video conferencing. Interviews lasted between 11 and 73 minutes (mean=33 minutes). Table 1 shows participants’ sociodemographic and clinical characteristics.

**Table 1.** Participant characteristics and sociodemographic data.

| Participant characteristics | Patients<br>(n = 8) | Informal caregivers<br>(n = 10) | Clinicians<br>(n = 10) |
| --- | --- | --- | --- |
| **Age category (years), n (%)** |  |  |  |
| 18 - 25 | - | 1 (10) | - |
| 26 - 35 | 1 (12.5) | 2 (20) | - |
| 36 - 45 | - | 2 (20) | 2 (20) |
| 46 - 55 | 2 (25) | 1 (10) | 5 (50) |
| 56 - 65 | 2 (25) | - | 2 (20) |
| 66 - 75 | 2 (25) | 4 (40) | - |
| 76 - 85 | 1 (12.5) | - | - |
| 85+ | - | - | - |
| Not reported | - | - | 1 (10) |
| **Gender, n (%)** |  |  |  |
| Male | 4 (50) | 4 (40) | 1 (10) |
| Female | 4 (50) | 6 (60) | 8 (80) |
| Prefer not to say | - | - | 1 (10) |
| **Ethnicity, n (%)** |  |  |  |
| Asian/Asian (British) | 1 (12.5) | - | - |
| Black (African) | 2 (25) | 1 (10) | - |
| Black (British) | - | 1 (10) | - |
| Mixed/Multiple | - | 2 (20) | - |
| White (British) | 2 (25) | 4 (40) | 9 (90) |
| White (Irish) | 1 (12.5) | - | - |
| White (other) | 2 (25) | 2 (20) | 1 (10) |
| **Patient specific characteristics** |  |  |  |
| **Patient type, n (%)** |  |  |  |
| Inpatient | 4 (50) | - | - |
| Outpatient | 4 (50) | - | - |
| **Care location, n (%)** |  |  |  |
| Hospice | 5 (62.5) | - | - |
| Hospital | 2 (25) | - | - |
| Community | 1 (12.5) | - | - |
| **Site of primary cancer, n (%)** |  |  |  |
| Liver | 1 (12.5) | - | - |
| Lung | 2 (25) | - | - |
| Lymphatic/haematological | 3 (37.5) | - | - |
| Other | 2 (25) | - | - |
| **Informal caregiver specific characteristics** |  |  |  |
| **Current caring situation, n (%)** |  |  |  |
| Bereaved caregiver | - | 6 (60) | - |
| Current caregiver | - | 4 (40) | - |
| **Relationship to patient, n (%)** |  |  |  |
| Child | - | 4 (40) | - |
| Friend | - | 1 (10) | - |
| Husband/Wife/Partner | - | 3 (30) | - |
| Other | - | 2 (20) | - |
| **Site of patient's primary cancer, n (%)** |  |  |  |
| Breast | - | 2 (20) | - |
| Liver | - | 1 (10) | - |
| Lung | - | 2 (20) | - |
| Lymphatic/haematological | - | 2 (20) | - |
| Pancreas | - | 1 (10) | - |
| Other | - | 2 (20) | - |
| **Clinician specific characteristics** |  |  |  |
| **Professional role, n (%)** |  |  |  |
| Physician | - | - | 7 (70) |
| Nurse | - | - | 3 (30) |
| **Role location, n (%)** |  |  |  |
| Hospital | - | - | 6 (60) |
| Hospice | - | - | 3 (30) |
| Community | - | - | 1 (10) |
| **Length of palliative care experience (years), n (%)** |  |  |  |
| 1 – 2 | - | - | 1 (10) |
| 2 – 5 | - | - | 1 (20) |
| 5 – 10 | - | - | 1 (10) |
| 10 – 15 | - | - | 1 (10) |
| 15 – 20 | - | - | 1 (10) |
| 20+ | - | - | 4 (40) |
| Not reported | - | - | 1 (10) |
| **Length of health care experience (years), n (%)** |  |  |  |
| 1 – 2 | - | - | - |
| 2 – 5 | - | - | - |
| 5 – 10 | - | - | - |
| 10 – 15 | - | - | 1 (10) |
| 15 – 20 | - | - | 2 (20) |
| 20+ | - | - | 6 (60) |
| Not reported | - | - | 1 (10) |

### Identification of outcomes and outcome domains

We identified 33 outcomes of prognostication from the interviews, 16 of which were unique to this study and not identified in the systematic review [9]. We categorised these outcomes into nine COMET domains, plus one additional domain identified in the literature review [9]: spiritual/religious/existential functioning/wellbeing. The resulting ten domains were: (1) mortality/survival; (2) general physiological/clinical outcomes; (3) psychiatric outcomes; (4) spiritual/religious/existential functioning/wellbeing; (5) emotional functioning/wellbeing; (6) social functioning; (7) delivery of care; (8) perceived health status; (9) personal circumstances; (10) societal/carer burden.

Table 2 details all outcomes from the interviews, organised by the COMET taxonomy outcome domains [10].

**Table 2.** Outcomes and outcome domains identified from the interview data.

| **Outcome domain** | **Outcomes** |
| --- | --- |
| Mortality/survival | Length of survival |
| Psychiatric outcomes | Depression |
|  | Anxiety |
| Physical outcomes | Pain |
| Spiritual/religious/existential functioning/wellbeing outcomes | Hopefulness/maintaining hope |
|  | Hopelessness/loss of hope |
|  | **Sense of control** |
|  | Wish to live |
|  | Desire for death |
|  | **Spiritual and religious coping** |
| Emotional functioning/wellbeing | **Disbelief, shock, and denial** |
|  | Emotional distress |
|  | **Avoidance of prognosis** |
|  | Prognostic acceptance |
|  | **Fixation on prognosis** |
|  | Having the opportunity to say goodbye to loved ones |
|  | **Mental/emotional preparation for EOL** |
|  | **Achieving/prioritising personal goals and values** |
|  | **Anticipatory grief** |
| Social functioning | **Quality of relationships with others** |
|  | **Quality of communication between patients and family/friends** |
| Delivery of care | **Shared decision-making** |
|  | **End-of-life/advance care planning** |
|  | Patient-doctor relationship |
|  | **Place of care** |
|  | **Access to practical support** |
|  | **Access to financial support** |
|  | Conflicting preferences for prognostic information between patients and caregivers |
|  | Dying in a preferred location |
| Perceived health status | Prognostic understanding |
|  | Being aware of prognostic uncertainty |
| Personal circumstances | Getting affairs in order |
| **Societal/carer burden** | **Caregiver/family burden** |
| New outcomes and domains which were not identified in the literature are presented in bold. |  |

#### Outcome domain 1: Mortality/survival

The outcome domain of ‘mortality/survival’ covers interviewees’ perspectives on how prognostication can impact patients’ length of survival. Both informal caregivers and clinicians conveyed how delivering a prognosis could heighten a patient’s awareness of their impending mortality, and so, unintentionally, impact their remaining lifespan:

> *“I found that after hearing that news my mum’s health declined. And I don’t think it was declining because of her illness. I think it declined because she feels like she’s dying because she’s just been told she is.” (Informal caregiver, IC03)*

#### Outcome domain 2: Physical outcomes

Clinicians were the sole group to identify physical outcomes, with a particular emphasis on the aspect of pain as a repercussion resulting from prognostic awareness. Clinicians underscored how prognostication might influence patients’ pain experiences, both positively and negatively:

> *“I saw her the following week and she’d just seen the consultant and been told-given a very specific prognosis of ten weeks … it definitely was influencing, I’m sure, yes, her pain, and her pain levels and her pain perception.” (Clinician, C02)*

> *“We talked about future, and it was a poor prognosis. And in the early days she was very pleased to have had the conversation; she felt unburdened by it. In fact, her pain improved quite a lot.” (Clinician, C07)*

#### Outcome domain 3: Psychiatric outcomes

Participants discussed the psychological burden faced by patients and their families when confronted with prognostication. The revelation of prognostic information might lead to heightened anxiety among patients and their families:

> *“I think my anxiety about the whole prognosis thing, I’m quite an anxious person but it’s now just like it’s a problem for me on a daily basis because just not knowing is just awful.” (Informal caregiver, IC10)*The psychological impact of prognostication also extended to depressive symptoms. Both clinicians and informal caregivers noted that receiving an unwelcome prognosis can precipitate depression or at least a significant lowering of mood. Patients themselves spoke to this experience:

> *“I mean I was for quite a while, quite depressed for a few-well, I’d say more than depressed.” (Patient, P04)*

#### Outcome domain 4: Spiritual/religious/existential functioning/wellbeing

From the interviews, it became evident that prognostication had multifaceted effects on the spiritual, religious, and existential aspects of individuals’ wellbeing. For some patients and informal caregivers, the knowledge of their prognosis brought about a tension or an alternation between hopefulness and hopelessness. Prognostication could inspire hope:

> *“…the doctor there was very open about it from quite early on. And he said to my partner to me, you know, jointly, this is very serious. It could be 50/50 … I thought well thinking of the 50 percent positive, not the 50 percent negative. There’s hope there.” (Informal caregiver, IC02)*A short-term prognosis could also lead to a palpable loss of hope, or hopelessness:

> *“I think my dad lost hope for a bit. So, once you kind of hear you are going to die soon, I do honestly, he knew he was ill but you don’t accept it, kind of you always hope you are going to live.” (Informal caregiver, IC09)*This tension in responses resonated with a broader sense of control. Stakeholders observed that prognostic information could either reinforce a sense of control, empowering individuals to make conscious decisions about their remaining time, or evoke a loss of control:

> *“For some people having the control and the knowledge, it’s beneficial for them, and to have lots of information early, and maybe as accurate information as can be given like a prognosis of three to six months is useful and they find it helpful for control … Whereas for someone else, actually I think they might get a prognosis and it can spin everything out of control for a while before they regroup.” (Clinician, C07)*Many patient participants identified spiritual and religious factors as important in facing their prognoses. They reported deriving comfort from their faith, or pursuing spiritual exploration as a means of finding peace and understanding, or completion:

> *“I’ve been visited by leaders of different religions … You spend your life asking if this is the right religion or this is the wrong religion. All you think is just your God, one God who loves us all. I am trying to find the answer before I go.” (Patient, P05)*Finally, the impact of prognostication on the will to live could also be polarised. Some participants indicated that a terminal prognosis could ignite a wish to live, but for others it could precipitate a desire for death:

> *“…since knowing my prognosis, I wake up and I know I cannot give up. I need to keep fighting this cancer and keep going. I want to keep living.” (Patient, P05)*

> *“Ultimately, she was asking about euthanasia and things like that, so I think having been given that short a prognosis, whatever that was based upon, had changed her outlook completely. She no longer wanted to live like that.” (Clinician, C02)*

#### Outcome domain 5: Emotional functioning/wellbeing

Emotional distress emerged as a prevalent emotional consequence of prognostication. Participants’ accounts revealed a complex array of emotional reactions. Initial responses were often disbelief, shock, and denial. This impact was evident in the personal accounts of patients:

> *“I was numb. I didn’t know. I said, “All right, thank you.” And that was it. I was just numb.” (Patient, P04)*Clinicians recognised substantial emotional reactions and adjustments in patients and their families, while informal caregivers and patients affirmed the considerable impact on their emotional state:

> *“Ever since they told me, my emotions are in turmoil.” (Patient, P03)*

> *“I just launched into a monologue about time and the prognosis … she slowly crumpled and started sobbing and pulled, literally pulled the bed clothes over her head and was in such distress…” (Clinician, C09)*Some attested that some people reacted to receiving a prognosis with avoidance, and defending against reality:

> *“I’ve had patients wave… very, very physically react and wave their hands in the air, when they say they don’t want to know, they’re actually waving their hands in front of their body as an almost… as a physical barrier, if they feel like you’re going to blurt it out to them…” (Clinician, C06)*Other participants reported finding acceptance, with patients shifting their focus to living fully or coming to terms with the inevitability of their situation:

> *“Obviously I felt scared and sad, but I said to myself, if it’s got to be, it’s got to be. There’s nothing else I can do, you know, unless a miracle happens. But miracles don’t happen” (Patient, P01)*Some clinician participants described scenarios where patients fixated on prognosis, often treating it as an absolute rather than an estimate. This fixation can have a considerable emotional impact:

> *“I’ve had patients who have taken what you’ve said as gospel, so I had a patient who I said, you know, short months, and they pushed me on that, and I said, ‘Maybe two or three.’ So, they literally ticked off days on a computer.” (Clinician, C01)*Some participants commented that prognostication can facilitate mental and emotional preparation for end-of-life, helping individuals and families to manage expectations and mentally acclimatise to the forthcoming changes:

> *“I think it’s-it can be very helpful for planning, and it can be helpful for loved ones to know, okay, this is what I need to prepare myself for, and also very much what might happen.” (Clinician, C03)*Some suggested that prognostication can facilitate the emotionally significant process of saying goodbye to loved ones:

> *“…for me that’s one of the benefits I have always thought of it [prognostication] is that you do, even if it’s just like a couple of days, you get that time to like say goodbye to everyone … it’s like one of the small silver linings in it I think” (Informal caregiver, IC08)*Others remarked that prognostication could act as a catalyst for individuals to focus on achieving or prioritising personal goals and values. Patients and their families often refocused their energy on what was most important to them, whether that be spending quality time with loved ones, fulfilling lifelong desires, or expediting important life events:

> *“Being told it was incurable, I was like, okay, that’s it now, the ticking clock has started now, if you know what I mean. So, we definitely did things quicker, as I said, we got married.” (Patient, P08)*Additionally, anticipatory grief was reflected in the experiences shared by informal caregivers and patients alike as they dealt with the emotional weight of a terminal prognosis:

> *“They [family] all scattered into just starting the grieving process immediately. And clearly, I’m not dying at the moment, you know.” (Patient, P02)*

#### Outcome domain 6: Social functioning

Participants’ comments indicated that prognostication could influence social functioning, by impacting the dynamics of relationships and communication. Participants in all three stakeholder groups recognised its role in either strengthening or reducing the quality of relationships with others:

> *“My children haven’t been in touch for a while, all of a sudden, they can’t stop calling me every day. My ex who I [was] hardly talking to but now they are staying with me pretty much every day, wishing me well.” (Patient, P05)*Some participants noted that the accuracy of prognostic information had an impact on communication between patients and their families and friends. One participant referred to “good quality” information, meaning precision:

> *“If somebody said weeks, months, or years, what do you do with that and how do you communicate? Oh my God I was sitting next to my partner, and she was ringing people to talk to them about it. And to be armed with good quality information would have made that process much easier.” (Informal caregiver, IC07)*

#### Outcome domain 7: Delivery of care

Participants revealed several pivotal aspects within the overarching domain of ‘delivery of care.’ Shared decision-making was frequently identified as an outcome, with participants from all three stakeholder groups emphasising the role of prognostication in guiding discussions, and ultimately informed choices, about treatment and care:

> *“It was my choice to stop maintenance therapy even though the doctor said patients who complete three years of maintenance therapy have X% longer … it’s a choice to make and you make it with the best available information.” (Patient, P07)*Participants also provided insight into how prognostication was intrinsically linked to end-of-life and advance care planning, often leading to considerations of ceilings of care, Do Not Resuscitate (DNR) orders, and other elements of end-of-life planning:

> *“I think if you’ve had the conversations about aims of treatment and outcomes and prognosis then that would then lead on to thinking about advanced care planning.” (Clinician, C02)*Some participants considered that the patient-doctor relationship was also affected by prognostication. Interviewees indicated that poor prognoses can lead to strained relationships and a loss of trust:

> *“I definitely have patients that we’ve come to see who have been given a poor prognosis and patients sort of feel like there’s a strained relationship because they feel like the team’s given up on them.” (Clinician, C08)*The significance of place of care was another focal point. Clinician participants underscored the value of prognostication in guiding discussions about care settings, whether in the hospice, hospital, or the community:

> *“I think it is hugely valuable for people to know their prognosis, not only in terms of where they might be looked after, not just being the hospice, but anywhere, hospital, community.” (Clinician, C04)*This included preferred location for dying, whether at home or in a medical facility:

> *“I think for some people, they might have a discussion about prognosis. If it was relatively short, they’d want to let everyone know there and then and say please write down everywhere that I don’t want to go back to hospital, I want to die at home.” (Clinician, C07)*Participants reflected on the practical and financial dimensions of care, with clinicians recognising the role of prognosis in facilitating access to essential support services, including referrals to hospice care, and fast-tracking community support:

> *“You need to know because we need to make sure that the support is there for you out in the community because you won’t be coming back to us in the hospital, so this is something we need to communicate to other people, but more importantly it’s something that they need to know.” (Clinician, C06)*A clear prognosis was also considered useful in guiding patients and their families to access financial benefits and allowances designed to assist them during challenging times:

> *“…essentially, I’ve retired early because I have limited time, so, I get a monthly income, a salary almost from it [pension]. I got a lump sum and so that helped us plan and do things.” (Patient, P08)*Our interviewees also highlighted the complex interplay of preferences for prognostic information within the patient-caregiver dyad. Clinicians and informal caregivers described scenarios where patients and family members held conflicting preferences for prognostic disclosure, underscoring the challenges faced by healthcare teams in navigating these social dynamics:

> *“Sometimes, you know, you’ll have a family member saying I don’t want you to tell her, I don’t want you to tell her, don’t you dare tell her … The patient is the person that is the centre, and the family is important but it’s actually secondary.” (Informal caregiver, IC01)*

#### Outcome domain 8: Perceived health status

Prognostic understanding was found to affect patients’ perception of their health status. Clinicians recognised the challenges in ensuring patients comprehend the uncertainty and limitations of prognostic information, aiming to reconcile patients’ understanding with the medical realities presented to them:

> *“I would broach it [prognostication] gently, kind of check understanding, double check understanding … it’s sort of like an iterative process, always reconfirming that they’re understanding what you’re saying…” (Clinician, C05)*The acknowledgement of prognostic uncertainty was also seen as important. Participants expressed a need for honesty and clarity, yet also recognised that precise predictions are not always possible:

> *“It must always come with that proviso that we don’t really know … that proviso must go in to any prognosis that we’re not really sure, you could live longer so that the patient knows.” (Informal caregiver, IC04)*

#### Outcome domain 9: Personal circumstances

‘Getting affairs in order’ was one of the most discussed issues during interviews with all three stakeholder groups. For informal caregivers and patients alike, knowing the prognosis served as a catalyst for arranging their personal, financial, and legal matters. The driving force behind these preparations often stemmed from a desire to alleviate potential burdens on themselves and loved ones. Clinicians acknowledged the importance of assisting individuals in addressing these practical aspects of their lives:

> *“I made sure that I also had enough money in my account to take care of me … I said to myself I don’t want to bring my children you know put [sic] debt for me. Like I don’t want them to struggle for money to bury me you know” (Patient, P06)*

> *“I think it can be, for some people it can be very helpful and especially, you know, if they’ve got family and they’ve got children or dependants, it kind of gives them those opportunities to put things in place for those.” (Clinician, C10)*

#### Outcome domain 10: Societal/carer burden

Comments from informal caregivers and clinicians underscored the societal and carer burden that can emerge when families are dealing with prognostic information.

Prognostication significantly impacted some informal caregivers’ lives, altering their personal and professional trajectories, and imposing new responsibilities that often led to substantial life changes:

> *“I was supposed to travel to Africa like tomorrow … But I can’t do that now because of this situation. So, I have to wait until whatever happens then I’ll know what, you know. So, it affects my movement, even work-wise, and everything. It affects everything about me right now.” (Informal caregiver, IC05)*

> *“Well, everything has changed since we found out [the prognosis]. My life has stopped. I became a full-time carer.” (Informal caregiver, IC06)*

## Discussion

This research aimed to identify outcomes associated with the impact of prognostication, from interviews with patients with advanced cancer, informal caregivers, and clinicians. [25]. Our analysis of the interview data identified 33 outcomes relevant for measuring the impact of prognostication. These outcomes underscore the complex nature of prognostication’s effects, spanning physical, emotional, social, psychological, and spiritual dimensions. Furthermore, they highlight how prognostication influences both patients and informal caregivers, carrying consequential implications for the delivery of healthcare.

Our study identified 16 outcomes that our previous literature review of prognostication studies did not find [9], highlighting a disjuncture between research and stakeholder priorities. This gap reinforces the need for outcome assessment methods that incorporate stakeholder perspectives. The need to fit outcome assessment to the evolving priorities of all key stakeholders is increasingly recognised in health and social care, and the implementation of validated COSs for people with conditions like rheumatoid arthritis and stroke exemplifies the advantages of this approach [26, 27]. Prognostication, too, could benefit significantly from a COS that respects and integrates the varied priorities of all stakeholders.

Our study also revealed distinct yet overlapping perspectives among stakeholders regarding important outcomes of prognostication. Frequently mentioned priorities across these groups included ‘achieving/prioritising personal goals and values,’ ‘shared decision-making’ and ‘getting affairs in order.’ This contrasts sharply with the lesser prioritisation of traditional clinical metrics, such as ‘pain’, which only our clinician participants mentioned. A more holistic view of prognostication mirrors broader trends in the field of palliative care, which has always emphasised holistic care, and where there is a growing consensus on the importance of addressing not just physical, but also psychological, social, and spiritual needs at end-of-life [28–30].

Understanding the experiences and perspectives of stakeholders has additional value, beyond informing the development of a COS. Some of the outcomes we identified may not be used in the final COS; however, their identification offers scope for refining and guiding future research. This might produce additional outcome measures, reflecting the priorities of patients, caregivers, and clinicians. For example, we identified ‘access to financial support’ as an important but previously under-recognised outcome, with particular relevance to recent discussions and campaigns focused on financial insecurity at end-of-life [31]. Such dimensions of prognostication can offer new insights into the quality of care, and highlight specific areas within healthcare and social policy which might be enhanced, such as advance care planning. Research and policy development addressing these additional outcomes might contribute to more comprehensive and effective end-of-life care provision, including considering how relevant these findings for people with advanced cancer are for people living with other terminal conditions.

### Strengths and limitations

To our knowledge, this is the first study to explore the experiences and perceptions of prognostication from the perspectives of patients, informal caregivers, and clinicians. A strength of our study is the diversity of participants, including stakeholders from various backgrounds, care settings, and with a range of advanced cancer diagnoses, so contributing to the broader applicability of our findings.

Purposive sampling was successful for many participant characteristics, but there was limited diversity in the clinicians’ gender and ethnicity. The predominance of female clinicians reflects the UK palliative medicine workforce’s demographics [32], but raises questions about potential gender-based differences in perspectives, which our study cannot address. Similarly, the limited ethnic diversity among clinicians could restricts multicultural understanding [33]. However, there was broad diversity in our patient and informal caregiver groups, which increases the relevance of our findings to multiple cultural contexts.

We recruited participants from a single UK region for pragmatic reasons, which limits the generalisability of these findings, due to variations in healthcare systems and cultural attitudes towards prognostication, nationally and internationally. However, the upcoming international Delphi survey for the final phase of the project will gather a wider array of perspectives, so broadening the applicability of the overall study.

## Conclusion

Our study identified 33 outcomes in ten domains that stakeholders from three key groups (patients, informal caregivers, and clinicians) deemed important to measure and report in future prognostic impact studies. These findings will inform the development of a comprehensive and relevant COS, and a more inclusive approach to outcome assessment, to better meet the diverse needs of patients, thereby enhancing the quality of palliative care. These findings may also pave the way for future research studies to explore under-recognised areas where prognostication is important.

## Data Availability

Data are available upon reasonable request. The data that support the findings of this study are available from the corresponding author upon reasonable request.

## Acknowledgements

We would like to thank all participants who kindly dedicated their time and insights to this study. We would also like to acknowledge the contribution of the palliative care teams at University College London Hospital, Central and North West London NHS Foundation Trust, and Marie Curie Hospice, Hampstead, who helped recruit participants for the study. Lastly, we extend our thanks to Marie Curie and the Economic and Social Research Council for supporting this study via a PhD Studentship grant.

## References

1. LeBlanc TW, Temel JS, Helft PR. “How Much Time Do I Have?”: Communicating Prognosis in the Era of Exceptional Responders. Am Soc Clin Oncol Educ Book. 2018;38:787–94. doi: 10.1200/edbk_201211. PubMed PMID: 30231384.

2. Owen R, Jeffrey D. Communication: common challenging scenarios in cancer care. Eur J Cancer. 2008;44(8):1163–8. Epub 20080324. doi: 10.1016/j.ejca.2008.02.029. PubMed PMID: 18364252.

3. Steyerberg EW, Moons KGM, van der Windt DA, Hayden JA, Perel P, Schroter S, et al. Prognosis Research Strategy (PROGRESS) 3: Prognostic Model Research. PLOS Medicine. 2013;10(2):e1001381. doi: 10.1371/journal.pmed.1001381.

4. Lambden J, Zhang B, Friedlander R, Prigerson HG. Accuracy of Oncologists’ Life-Expectancy Estimates Recalled by Their Advanced Cancer Patients: Correlates and Outcomes. J Palliat Med. 2016;19(12):1296–303. Epub 2016/08/31. doi: 10.1089/jpm.2016.0121. PubMed PMID: 27574869; PubMed Central PMCID: PMCPMC5144872.

5. Farinholt P, Park M, Guo Y, Bruera E, Hui D. A Comparison of the Accuracy of Clinician Prediction of Survival Versus the Palliative Prognostic Index. Journal of Pain and Symptom Management. 2018;55(3):792–7. doi: 10.1016/j.jpainsymman.2017.11.028.

6. Seung Hun L, Jeong Gyu L, Young Jin C, Young Mi S, Hyojeong K, Yun Jin K, et al. Prognosis palliative care study, palliative prognostic index, palliative prognostic score and objective prognostic score in advanced cancer: a prospective comparison. BMJ Support Palliat Care. 2021:bmjspcare-2021-003077. doi: 10.1136/bmjspcare-2021-003077.

7. Baba M, Maeda I, Morita T, Inoue S, Ikenaga M, Matsumoto Y, et al. Survival prediction for advanced cancer patients in the real world: A comparison of the Palliative Prognostic Score, Delirium-Palliative Prognostic Score, Palliative Prognostic Index and modified Prognosis in Palliative Care Study predictor model. European Journal of Cancer. 2015;51(12):1618–29. doi: 10.1016/j.ejca.2015.04.025.

8. Todd S, David CM, Karel GMM, Amanda JC, Mattias J, Pietro F, et al. Comparison of prognostic models to predict the occurrence of colorectal cancer in asymptomatic individuals: a systematic literature review and external validation in the EPIC and UK Biobank prospective cohort studies. Gut. 2019;68(4):672. doi: 10.1136/gutjnl-2017-315730.

9. Spooner C, Vivat B, White N, Bruun A, Rohde G, Kwek PX, et al. What outcomes do studies use to measure the impact of prognostication on people with advanced cancer? Findings from a systematic review of quantitative and qualitative studies. Palliative Medicine. 0(0):02692163231191148. doi: 10.1177/02692163231191148.

10. Dodd S, Clarke M, Becker L, Mavergames C, Fish R, Williamson PR. A taxonomy has been developed for outcomes in medical research to help improve knowledge discovery. J Clin Epidemiol. 2018;96:84–92. Epub 2017/12/31. doi: 10.1016/j.jclinepi.2017.12.020. PubMed PMID: 29288712; PubMed Central PMCID: PMCPMC5854263.

11. Spooner C, Vivat B, White N, Stone P. Developing a Core Outcome Set for Prognostic Research in Palliative Cancer Care: Protocol for a Mixed Methods Study. JMIR Res Protoc. 2023;12:e49774. Epub 20230901. doi: 10.2196/49774. PubMed PMID: 37656505; PubMed Central PMCID: PMCPMC10504625.

12. Glare P, Virik K, Jones M, Hudson M, Eychmuller S, Simes J, et al. A systematic review of physicians’ survival predictions in terminally ill cancer patients. BMJ. 2003;327(7408):195. doi: 10.1136/bmj.327.7408.195.

13. White N, Reid F, Harris A, Harries P, Stone P. A Systematic Review of Predictions of Survival in Palliative Care: How Accurate Are Clinicians and Who Are the Experts? PLOS ONE. 2016;11(8):e0161407. doi: 10.1371/journal.pone.0161407.

14. White N, Kupeli N, Vickerstaff V, Stone P. How accurate is the ‘Surprise Question’ at identifying patients at the end of life? A systematic review and meta-analysis. BMC Med. 2017;15(1):139. Epub 2017/08/03. doi: 10.1186/s12916-017-0907-4. PubMed PMID: 28764757; PubMed Central PMCID: PMCPMC5540432.

15. Coulter A. Measuring what matters to patients. BMJ. 2017;356:j816. doi: 10.1136/bmj.j816.

16. Williamson PR, Altman DG, Bagley H, Barnes KL, Blazeby JM, Brookes ST, et al. The COMET Handbook: version 1.0. Trials. 2017;18(Suppl 3):280. Epub 2017/07/07. doi: 10.1186/s13063-017-1978-4. PubMed PMID: 28681707; PubMed Central PMCID: PMCPMC5499094.

17. Williamson PR, Altman DG, Blazeby JM, Clarke M, Devane D, Gargon E, et al. Developing core outcome sets for clinical trials: issues to consider. Trials. 2012;13:132. Epub 2012/08/08. doi: 10.1186/1745-6215-13-132. PubMed PMID: 22867278; PubMed Central PMCID: PMCPMC3472231.

18. Bunniss S, Kelly DR. Research paradigms in medical education research. Medical education. 2010;44(4):358–66.

19. Tong A, Sainsbury P, Craig J. Consolidated criteria for reporting qualitative research (COREQ): a 32-item checklist for interviews and focus groups. International Journal for Quality in Health Care. 2007;19(6):349–57. doi: 10.1093/intqhc/mzm042.

20. Hennink MM, Kaiser BN, Marconi VC. Code Saturation Versus Meaning Saturation: How Many Interviews Are Enough? Qual Health Res. 2017;27(4):591–608. Epub 20160926. doi: 10.1177/1049732316665344. PubMed PMID: 27670770; PubMed Central PMCID: PMCPMC9359070.

21. Saunders B, Sim J, Kingstone T, Baker S, Waterfield J, Bartlam B, et al. Saturation in qualitative research: exploring its conceptualization and operationalization. Qual Quant. 2018;52(4):1893–907. Epub 20170914. doi: 10.1007/s11135-017-0574-8. PubMed PMID: 29937585; PubMed Central PMCID: PMCPMC5993836.

22. QSR International Pty Ltd. NVivo (Version 12) Chadstone, VIC, Australia: QSR International Pty Ltd; 2018 [cited 2023 12 July]. Available from: https://www.qsrinternational.com/nvivo-qualitative-data-analysis-software/home?_ga=2.10629428.1253174092.1679915032-931267505.1679915032.

23. Gale NK, Heath G, Cameron E, Rashid S, Redwood S. Using the framework method for the analysis of qualitative data in multi-disciplinary health research. BMC Medical Research Methodology. 2013;13(1):117. doi: 10.1186/1471-2288-13-117.

24. Ritchie J, Lewis J, Nicholls CM, Ormston R. Qualitative research practice: A guide for social science students and researchers: Sage; 2013.

25. Keeley T, Williamson P, Callery P, Jones L, Mathers J, Jones J, et al. The use of qualitative methods to inform Delphi surveys in core outcome set development. Trials. 2016;17:1–9.

26. Budhdeo S, Chari A, Harrison O, Blazeby J. Patient-centred healthcare outcome measures: towards a unified architecture. J R Soc Med. 2014;107(8):300–2. Epub 20140801. doi: 10.1177/0141076814545701. PubMed PMID: 25086055; PubMed Central PMCID: PMCPMC4128084.

27. Gargon E, Gurung B, Medley N, Altman DG, Blazeby JM, Clarke M, et al. Choosing Important Health Outcomes for Comparative Effectiveness Research: A Systematic Review. PLOS ONE. 2014;9(6):e99111. doi: 10.1371/journal.pone.0099111.

28. Clark D. From margins to centre: a review of the history of palliative care in cancer. The Lancet Oncology. 2007;8(5):430–8. doi: 10.1016/S1470-2045(07)70138-9.

29. Clark D. ’Total pain’, disciplinary power and the body in the work of Cicely Saunders, 1958-1967. Soc Sci Med. 1999;49(6):727–36. doi: 10.1016/s0277-9536(99)00098-2. PubMed PMID: 10459885.

30. Saunders C, Summers DH, Teller N. Hospice : the living idea. London: Edward Arnold; 1981.

31. Marie Curie. Dying in Poverty: Improving financial support for terminally ill people with the cost of living 2022 [cited 2023 15 Nov]. Available from: https://www.mariecurie.org.uk/globalassets/media/documents/policy/policy-publications/2022/marie-curie-dying-in-poverty-budget-briefing-2022.pdf.

32. The Association for Palliative Medicine (APM) of Great Britain and Ireland. Report and overview of the pallaitive medicine workforce in the United Kingdom 2019 [cited 2023 15 Nov]. Available from: https://apmonline.org/wp-content/uploads/2019/08/palliative-medicine-workforce-report-2019-2.pdf.

33. Heena K, Derek W. Lack of racial diversity within the palliative medicine workforce: does it affect our patients? BMJ Support Palliat Care. 2022;12(1):49. doi: 10.1136/bmjspcare-2021-003117.

