## Supplemental material 1 for "Outcomes of prognostication in people living with advanced cancer: a qualitative study to inform a Core Outcome Set" (from another version of this preprint: version 1, 2024-02-08)

**PATIENT INTERVIEW GUIDE**

**Establishing Rapport**

To begin with, would you tell me a little bit about why you were interested in taking part in this study?

**Semi-structured interview guide**

1. Can you tell me how you were first told about your prognosis?

**Prompts:**

- By whom?
- Was anyone else present with you at the time (caregiver)?

2. When did this happen? How long ago?

**Prompts:**

- If you cannot remember specific date, perhaps just the year?

3. Would you rather have been told this sooner or later than actually happened?

**Prompts:**

And why is that?

4. Did you find it helpful or unhelpful knowing your likely prognosis?

**Prompts:**

In what ways?

How did knowing your prognosis affect your feelings?

How did knowing your prognosis affect your relationship with your family/friends/informal caregivers?

How did knowing your prognosis affect how you made plans?

How did knowing your prognosis affect you financially?

5. Do you think this would have affected any aspect of your care?

**Prompts:**

What? How?

How did knowing your prognosis affect choices and decisions you made about your care?

6. What kind of effect on your lives do you think would it have had if you had known sooner/later?

**Prompts:**

How? Why?

7. Is there anything that you would have done differently if you didn’t know your prognosis?

**Prompts:**

What? Why?

8. What information do you think is important for a palliative care clinician to tell patients and caregivers about their prognosis?

**Prompts:**

- There are different ways that people can be told about the likely prognosis. Some doctors use very broad general terms, like good or not so good; others talk about probabilities, like 40% chance. Some are a bit more specific, like days or weeks, or some will be as specific as they can be, and tell their patients the length of time they expect, like three months or five or six weeks. What do you think about these approaches? Do you think any of them are better or worse?
- Are there any other details that matter?
- Why are these things important?

9. How do you think patients and their caregivers should be told about the likely prognosis?

**Prompts:**

- How? Why?
- Would you like information to be available online, or only provided in person by a professional?
- Who should start talking about prognosis? The patient or the doctor or nurse?

10. Which of the things (or *effects*) that you have mentioned do you consider the most important?

**Prompts:**

- Which has affected you the most?
- Why is/was this important?

**Conclusion**

11. This is my last question. Are there any other good or bad things about knowing your prognosis that you can think of that we haven't discussed today?

**CAREGIVER INTERVIEW GUIDE**

**Establishing Rapport**

Before we begin, it would be helpful if you could tell me a little bit about why you were interested in taking part in this study.

**Semi-structured interview guide**

1. Were you present when [care recipient] was told their prognosis?

**Prompts:**

- *If YES:* By whom?
- *If NO:* When did you first hear about [care recipient's] prognosis? Who told you? When?

2. When did this happen? How long ago?

**Prompts:**

- If you cannot remember specific date, perhaps just the year?

3. Would you rather have been told the prognosis sooner or later than actually happened?

**Prompts:**

- And why is that?

4. Did you find it helpful or unhelpful knowing [care recipient’s] likely prognosis?

**Prompts:**

- In what ways?
- How did knowing their prognosis affect your feelings
- How did knowing their prognosis affect your relationship with them?
- How did knowing their prognosis affect how you made plans?
- How did knowing their prognosis affect you financially?

5. Do you think this would have affected any aspect of [care recipient’s] care?

**Prompts:**

- What? How?
- How did knowing their prognosis affect choices and decisions made about their care?

6. What kind of effect on your lives do you think would it have had if you had known sooner/later?

**Prompts:**

How? Why?

7. Is there anything you would have done differently if you didn’t know their prognosis?

**Prompts:**

- What? Why?

8. What information do you think is important for a palliative care clinician to tell patients and caregivers about their prognosis?

**Prompts:**

- There are different ways that people can be told about the likely prognosis. Some doctors use very broad general terms, like good or not so good; others talk about probabilities, like 40% chance. Some are a bit more specific, like days or weeks, or some will be as specific as they can be, and tell their patients the length of time they expect, like three months or five or six weeks. What do you think about these approaches? Do you think any of them are better or worse?
- Are there any other details that matter?
- Why are these things important?

9. How do you think patients and their caregivers should be told about the likely prognosis?

**Prompts:**

- How? Why?
- Would you like information to be available online, or only provided in person by a professional?
- Who should start talking about prognosis? The patient or the doctor or nurse?

10. Which of the things (or *effects*) that you have mentioned do you consider the most important?

**Prompts:**

- Which has affected you the most?
- Why is/was this important?

**Conclusion**

11. This is my last question. Are there any other good or bad things about knowing [care recipient’s] prognosis that you can think of that we haven't discussed today?

**CLINICIAN INTERVIEW GUIDE**

**Establishing Rapport**

Before we begin, it would be helpful if you could tell me a little bit about why you were interested in taking part in this study.

**Semi-structured interview guide**

1. How do you usually approach prognostic conversations?

**Prompts:**

- Do you use a specific algorithm/method to deliver a prognosis?
- What information do you think patients and their caregivers need to know about their prognosis?
- Do you use any resources to assist them in understanding their prognosis? For example, pamphlets, pictures, etc.? If not, why not? If so, how useful, or not useful do you think they were in helping them to understand their prognosis?
- Do you think it’s better to provide estimates in terms of days, weeks, or months, or as probabilities, or using general broad terms such as good or not so good?
- Do you refer patients and their caregivers to further support? What/who?

2. In your experience, do you think that providing a prognosis is helpful or unhelpful to patients and their caregivers?

**Prompts:**

- In what ways?
- How do you think knowing their prognosis might affect how they make plans?
- How do you think knowing their prognosis might affect them financially?

3. In your experience, do you think that providing a prognosis is helpful or unhelpful clinically?

**Prompts:**

- In what ways?
- For example, how do you think knowing a patient's prognosis might help with planning care?
- What about with prioritising caseload?
- What about admission and discharge decisions?
- What about communication with colleagues?
- What about referring patients to hospice services?

4. You probably find that patients and their families or friends are upset after [you/their clinician] have shared their prognosis with them. Do you notice any other changes in patients?

**Prompts:**

- For example, does their physical health improve, decline, or stay the same? Can you provide an example of that?
- What about their mental health?
- What about their relationships?
- What do you think might be the reasons for these changes?

5. Do you think providing a prognosis might affect any particular aspect of a patient’s care?

**Prompts:**

- What? How?

6. Do you measure any outcomes or effects of providing prognosis to patients and their caregivers?

**Prompts:**

- If yes, how?
- If no, do you have any thoughts on how we might measure any effects or outcomes of providing prognoses?

7. Which of the things (or *effects*) that you have mentioned do you consider the most important?

**Prompts:**

- Which do you think would affect patients and/or caregivers the most?
- Why is/was this important?

**Conclusion**

8. This is my last question. Are there any other good or bad things about providing a prognosis that you can think of that we haven't discussed today?

__________________________________________________________________________________

**End**

Thank you very much for your time and the information you shared today.

Would you like to be kept informed of the results of my study? If so, are you happy for me to keep your contact details?

Non-leading and general prompts:

- Can you tell me more about that?
- Tell me what that was like for you?
- Why is that?
- Can you clarify more about [insert]?
- You mentioned [insert], can you describe what you mean by that?
- Reflect on answer, summarise to check, and prompt
